# The Relationship Between Electrodermal Activity and Cardiac Troponin in Patients with Paroxysmal Sympathetic Hyperactivity

**DOI:** 10.1101/2024.09.14.24313588

**Authors:** Louis Beers, Jana Bouvain, Claus Reinsberger, Rasmus Jakobsmeyer, Rani Sarkis, Jong Woo Lee

## Abstract

**Objective:** To determine the relationship between electrodermal activity (EDA) and cardiac troponin in patients with paroxysmal sympathetic hyperactivity (PSH).

**Methods:** This was a study with prospectively-identified patients and a retrospective analysis utilizing electrodermal data taken from the wrist-worn Empatica E4 device (Empatica Srl, Milan, Italy) and troponin values obtained from critically-ill patients with suspected PSH (N=10). The maximum EDA value and temporally-nearest cardiac troponin were correlated to test for significance using Pearson correlation coefficient.

**Results:** A moderate correlation was found between EDA and troponin in 10 patients using the most temporally-proximal troponin to each patient’s maximal EDA (r=0.634; p = 0.049). A subanalysis was performed excluding any patients whose available troponin data did not fall within seven days of their maximal EDA, which demonstrated a strong correlation (r=0.943; p = 0.005). No relationships between troponin and pulse, blood pressure, or temperature were found.

**Conclusions:** This study establishes an association between wrist-worn EDA and a measure of possible myocardial injury in critically ill patients with PSH. Patients with elevated sympathetic activity may be at increased risk for concurrent cardiac injury or dysfunction, and thus EDA data from wrist-worn monitors may provide clinically significant data regarding sympathetic function.

## Introduction

Paroxysmal sympathetic hyperactivity (PSH), also known as sympathetic storming, describes sudden, reoccurring increases in autonomic activity, often caused by brain injury. It is a distinct form of dysautonomia in which there is an increase in sympathetic nervous system activity. [1] Certain physiological symptoms will often accompany these events, some of which are an increased heart rate, systolic blood pressure, temperature, respiratory rate, and diaphoresis. [2,3]

The most widely accepted understanding of why this occurs is known as the excitatory: inhibitory ratio (EIR) model. It is thought that upon damage to the brain such as a traumatic brain injury (TBI) or stroke, a lesion forms that disrupts the excitatory and inhibitory centers in the brainstem or spinal levels. This disruption may consequently lead to an impaired inhibition resulting in an increased excitation (sympathetic storming). [4]

Electrodermal activity (EDA) is used for measuring pure sympathetic activity, as parasympathetic input is excluded. EDA measures the conductance at the skin surface via sweat gland activity. [5] Because of this, it is possible to use wireless sensors non-invasively in order to determine when a patient is experiencing sudden increases in sympathetic activity. [6] As EDA offers the only current means of measuring pure sympathetic tone, it can be used in a clinical setting to detect paroxysmal sympathetic hyperactivity. Identifying cases of PSH is clinically significant since patients who exhibit symptoms often have a less favorable outcome. On average, patients with PSH will have longer hospital stays, worse Disability Rating Scale (DRS) scores, and higher death rates. [7]

Sympathetic storms may place an additional burden on cardiac function. Often with myocardial injury, there will be a release of troponin into the bloodstream. [8] This study sought to determine if there exists a correlation between sympathetic tone, as measured by EDA, and cardiac troponin, which would indicate a possible relation between PSH and cardiac injury.

## Materials and Methods

### Patients

This study included prospectively-identified patients who were admitted to Brigham and Women’s Hospital between March 2016 to September 2018. Patients were recruited from several ICUs, including neuroscience and surgical ICUs, and referred for EDA monitoring when they had an underlying neurological diagnosis and there was clinical concern for sympathetic hyperactivity based on tachycardia, tachypnea, hypertension, hyperthermia, hyperhidrosis, or posturing. [9] As patients were identified due to clinical concern, no strict clinical criteria were utilized to exclude patients.

The Institutional Review Board approved this study and all patients consented to the study. The study was also in compliance with the 2013 version of the Declaration of Helsinki. Inclusion criteria consisted of patients with clinically expected paroxysmal sympathetic hyperactivity based on their history and clinical presentation. Exclusion criteria consisted of patients who were unable to wear the EDA recording device, generally due to arterial line placement over the region of device placement and skin breakdown. De-identified data will be available upon request.

This was a retrospective analysis of patients with both documented troponin levels and prospectively-collected EDA data. EDA recordings were visually inspected by a board-certified physician adept in reading EDA signals to identify recordings exhibiting sympathetic hyperactivity. Recordings were continuous, with an average length of 18 hours 40 minutes 47 seconds and standard deviation of 6 hours 9 minutes 58 seconds. Empatica had no role in data analysis. Troponins were collected as part of the patients’ clinical care, as deemed appropriate, where there was concern for myocardial injury. As risk of myocardial injury was not always felt to be present despite concerns for PSH, troponin levels were not uniformly drawn. The patients consented for EDA measurement and medical record analysis only. This was not part of a bigger trial but a pilot study of wrist-worn EDA in the ICU.

The most proximal troponin value to the maximum EDA recording was collected. After identifying and measuring maximum EDA peak and corresponding troponin values, a Pearson correlation with p-value test was analyzed to test for a significant relationship. To further investigate a temporal relationship between EDA and troponin, a subanalysis was performed excluding patients whose troponin was not measured within seven days of their maximal EDA recording.

### Device

The Empatica E4 wristband (Empatica Srl, Milan, Italy) was used to measure EDA in patients with sympathetic storms. The device records EDA, blood volume pulse, accelerometer, heart rate calculated from inter-beat intervals, and skin temperature. EDA is collected using two electrodes that are placed on the ventral wrist to measure fluctuating changes in the electrical output of the skin. [10] Raw electrodermal data is collected at a sample rate of 4 Hz with values reported in microsiemens (µS). The range of detectable EDA output is from 0.01-100 µS. The EDA sensor also features a resolution of ∼900 pS and an alternating current at 8 Hz with a maximum peak to a peak value of 100 µAmps. [11] Data are transferred to a cloud-based system on the Empatica website (E4 connect), where they may be analyzed visually or compiled into a spreadsheet containing raw data.

## Results

In total, there were 120 EDA recordings across 39 patients. A sample recording is shown in Figure 1. From the board-certified physician’s analysis, it was determined that 54 recordings across 22 patients demonstrated interpretable artifact-free signals. The maximum EDA value for each of these patients was recorded. Of these 22 patients, 21 had identifying information recorded, and the characteristics of this group are shown in Table 1. A total of 10 of these 21 patients had troponin data available and every value during their hospital stay was recorded. Figure 2 denotes patient exclusions to reach a final group of 10 patients.

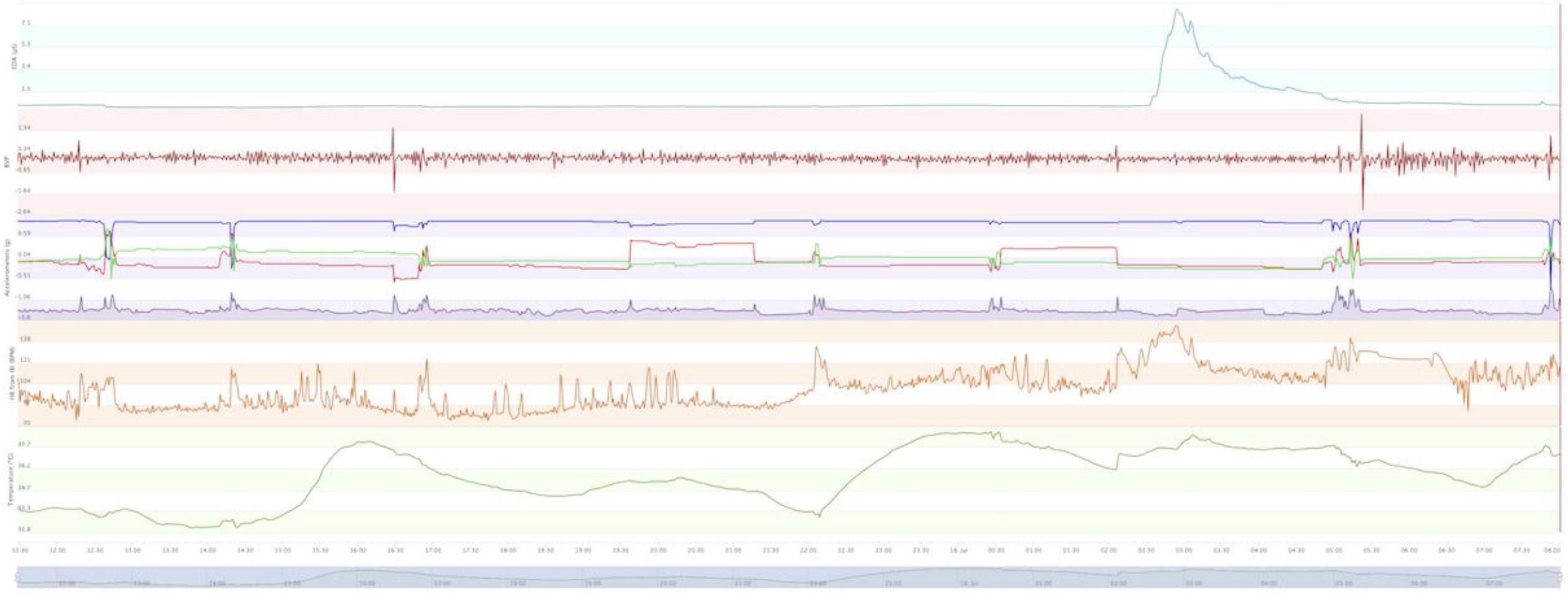

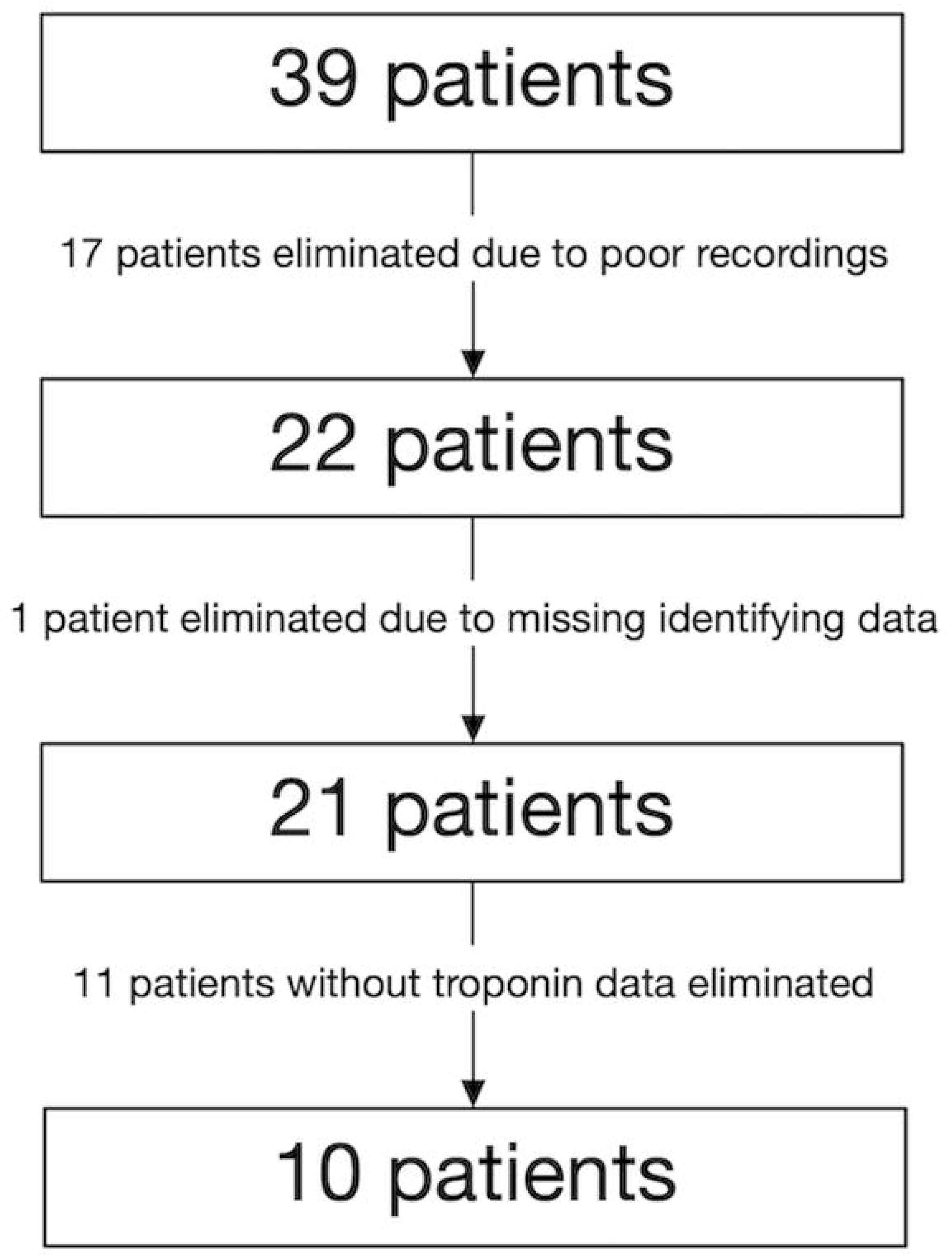

|  |  |
| --- | --- |
| Number of patients | 10 |
| Demographics |  |
| White | 10 (100.0%) |
| Sex Assigned at Birth |  |
| Male | 4 (40.0%) |
| Female | 6 (60.0%) |
| Age at Presentation (years) |  |
| Range | 28 – 73 |
| Average (± SD) | 51.5 ± 13.9 |
| Underlying neurological condition |  |
| Intracranial hemorrhage | 5 (50.0%) |
| Guillain-Barré syndrome or variant | 2 (20.0%) |
| Stroke | 1 (10.0%) |
| Chordoma | 1 (10.0%) |
| Encephalomyelitis | 1 (10.0%) |

The 10 patients with artifact-free EDA surges and troponin recordings were further analyzed. EDA values ranged from 3.53 to 90.84 µS. There was a statistically significant correlation between EDA and troponin values, with a Pearson correlation coefficient of 0.634 (Figure 3, *p* = 0.049). To address possible concerns of a lack of temporal relationship between troponin and EDA, a subanalysis of 6 patients was performed, excluding patients who did not have a recorded troponin level within seven days of their maximal EDA value. The average time between troponin and EDA in the subanalysis was 1 day, 12 hours, 42 minutes, and 50 seconds with a standard deviation of 23 hours, 47 minutes, and 30 seconds. Subanalysis EDA values of patients with troponin levels within 7 days ranged from 6.17 to 71.02. A statistically significant correlation was found between EDA and troponin in this subanalysis with a Pearson correlation coefficient of 0.943 (*p* = 0.005). EDA was also correlated with heart rate, blood pressure, and temperature, but there were no significant findings.

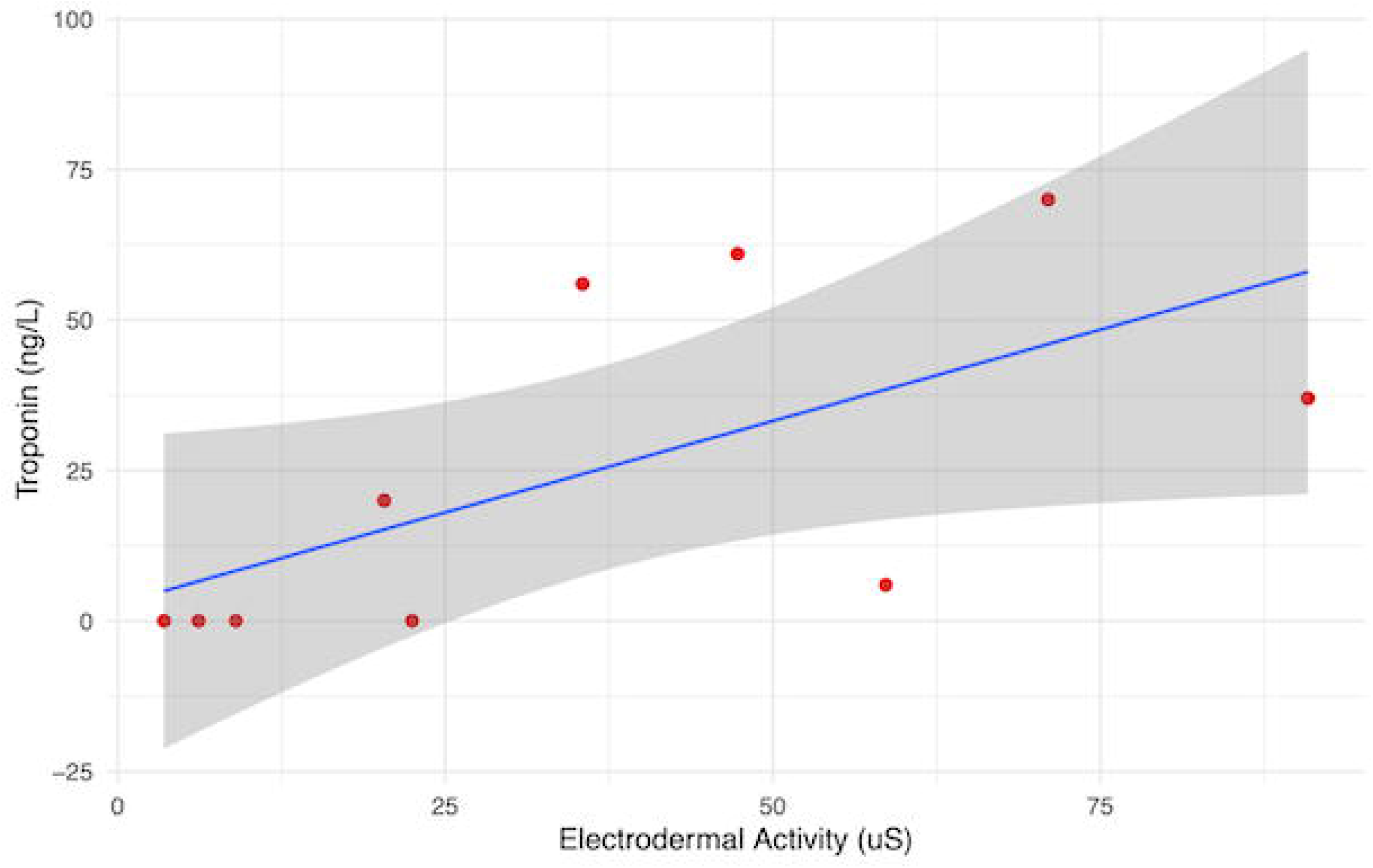

## Discussion

In this prospective study of patients with elevated electrodermal activity indicative of paroxysmal sympathetic hyperactivity, we found that maximal elevated EDA recordings were associated with raised troponin values. Since elevated cardiac troponin levels in the bloodstream are often indicative of myocardial injury, this suggests an associative relationship between sympathetic hyperactivity with cardiac injury. We demonstrate that neurophysiological biomarkers correlate to serum markers, which, to our knowledge, had not previously been demonstrated.

Previous studies have demonstrated that sympathetic storms can lead to poor cardiac outcomes. A prominent example of this is Takotsubo cardiomyopathy (TCM), otherwise known as stress-induced cardiomyopathy, in which there is a transient ballooning of the left ventricular apical segment after physical or emotional stress and an increase in plasma cardiac troponin levels. [12,13] Patients with TCM have symptoms similar to an acute coronary syndrome, and are at a greater risk for adverse events, complications, and mortality, with a 15-year mortality risk similar to patients with myocardial infarction. [14,15] A proposed mechanism for the pathophysiology of TCM involves epinephrine, indicating a link between sympathetic storms and cardiac dysfunction. [16] Chronic sympathetic hyperactivity may also lead to poor cardiac outcomes, as seen in hypertrophic cardiomyopathy. Animal studies have demonstrated that elevated levels of plasma catecholamines may lead to cardiac hypertrophy. [17,18] Furthermore, strenuous physical activity can lead to elevated troponin levels. [19] The physiological symptoms during sympathetic storms may place a similar stress on the heart, leading to troponin leaks.

EDA recordings are a unique way of quantifying a patient’s pure sympathetic activity, utilizing skin conductance which is unaffected by parasympathetic output, unlike other physiological measures typically used to assess sympathetic hyperactivity such as pulse, blood pressure, and temperature. A non-invasive EDA recording offers a means to determine a patient’s sympathetic output at any given time and data may be easily visualized to determine paroxysmal sympathetic hyperactivity. We chose to find the maximum EDA value and the most temporally proximal cardiac troponin plasma level during patients’ hospital stays, as this was most representative of the relationship between sympathetic hyperactivity and plasma troponin levels. As much is unknown about the exact mechanism between sympathetic storming and cardiac troponin release, such as if there exists an EDA threshold that must be reached for troponin to be released, we compared maximum EDA and most proximal troponin, as this methodology would avoid such issues. Of note, we did not find a relationship between troponin and either heart rate, blood pressure, or skin temperature, which may indicate the advantage of using EDA to track sympathetic hyperactivity, as it is not under any parasympathetic control.

With cardiac troponins being a prominent biomarker of cardiac injury, it is of clinical importance to identify the causes of their release into the bloodstream. Once identified, treatment of the underlying cause may result in a decrease in cardiac stress. Our finding of a correlation between electrodermal activity and troponin levels indicates a promising future in reducing cardiac issues after paroxysmal sympathetic hyperactivity. In TCM, patients are at an increased risk of morbidity and mortality during, and especially after, the acute phase of the disease. However, treatment with angiotensin-converting-enzyme inhibitors or angiotensin-receptor blockers improves survival. [14] This indicates that controlling sympathetic symptoms in patients with paroxysmal sympathetic hyperactivity after neurological dysfunction may also lead to improved cardiac outcomes. As PSH is most frequently seen after traumatic brain injury with hemorrhage, patients presenting with a TBI or other neurological condition causing brain injury may benefit from early monitoring of sympathetic tone. A wrist-worn device, such as the Empatica E4 wristband offers a simple, non-invasive method to continuously record EDA, allowing clinicians to monitor spikes in sympathetic activity. If there were to be a sympathetic storm indicated by the EDA, controlling symptoms may prevent cardiac dysfunction. Further studies are required to determine whether optimal treatment of sympathetic surges detected by EDA has the potential for improving cardiac outcomes in these patients.

There are several strengths to this study. With the usage of the E4 wristband, patients’ EDAs are measured from a common source, decreasing the risk of deviations from differing collection methods. Additionally, once the wristband is secured, data collection is standardized and quantifiable results are given, mitigating the risk of provider bias. Troponin levels were all taken at the same institution. This also leads to a low risk of deviation between data collection methods and provider bias. The inclusion of a subanalysis with more temporally-proximal recordings indicating an even stronger association between EDA and troponin also presents a strength to this study, as it further supports and strengthens the primary conclusion that there may be a relation between the two.

There are several study limitations. After excluding patients such that only those with artifact-free EDA recordings and recorded troponin values remained, there were only 10 subjects for analysis. Most of these artifacts occurred due to technical difficulties with the device or operator error. With a limited sample size, the significance of the conclusions is relatively modest. Additionally, some patients were already treated for their sympathetic symptoms which could have led to a decrease in the strength of association between EDA and troponin levels. The remaining population was heterogeneous in terms of etiology of PSH. However, the mechanism of the study remains consistent among participants as all had sympathetic storms. The interpretability of the EDA recordings was performed by visual inspection which may also create some degree of subjectivity in the dataset. An additional confounder was that troponins were not taken in every patient; they were more likely to be taken in those with a higher suspicion of cardiac disease. Even though results were potentially stratified from a high-risk group, the relationship between troponin and EDA levels remained, and inclusion of low-risk groups likely would have resulted in a stronger association. Bilateral recordings were often not obtainable due to the presence of arterial lines in many patients, despite findings that electrodermal activity may be asymmetrical. [20]

Future steps may be taken to avoid the technical difficulties and operator error that occurred during data collection, allowing for more robust and reliable recordings. Technical improvement to the device itself may provide an avenue for improvement in data collection. On multiple occasions, data was lost due to failure of the device to communicate leading to a reduction in the number of recordings. Modifying the device further to allow for an improved wristband fit may also be of benefit. Patient populations typically include a wide array body shapes and many patients, particularly those with concern for cardiac dysfunction, may be edematous as well. As such, multiple recordings were collected in patients in which the wrist-watch fit tightly, which may not have produced ideal data. In these patients, utilizing alternative devices placed on the palm or finger may be explored, as this could yield recordings of improved quality, with the disadvantage that such devices are more likely to detach during routine care.

## Conclusions

There is a significant relationship between maximum EDA values, as measured by a non-invasive wrist-worn device, and proximal cardiac troponin levels. These findings indicate that patients with increased sympathetic activity may be at increased risk for cardiac dysfunction. Sympathetic tone may be measured with wrist-worn EDA monitors, although other skin conductance modalities may be explored so that patients unable to wear a wrist sensor (e.g. if arterial line placed) may have their sympathetic activity measured. Further studies are required to determine whether management based on the measured sympathetic tone post-injury results in improvement of cardiac dysfunction.

## Data Availability

De-identified data will be available upon request.

